# Long Covid in Older Adults: Functional Outcomes and Treatment Effectiveness

**DOI:** 10.1101/2025.01.31.25321501

**Authors:** Brittin Wagner, Kate Mathers, Aaron Norfolk, Tanishtha Arora

**Author notes:** PointClickCare, 3500 American Blvd. W. Suite 150., Bloomington, MN 55431. All authors are employees of PointClickCare Technologies with no competing interests to declare.

## Abstract

We describe six-month functional status, hospitalization, and mortality among 23,632 cases of Long Covid in skilled nursing facilities (SNFs) in the US, primarily experienced by adults over 65 years old. We describe outcomes across seven medications used to treat Covid-19 including functional status (ADLs).

## Understanding Long Covid in Older Adults in Long-term Care

Understanding Long Covid in older adults poses distinct difficulties due to its diverse symptoms and the absence of targeted diagnostic tests. Long Covid is a condition characterized by symptoms that persist or develop for up to four weeks or more after a Covid-19 infection [2] and is recognized as a billable diagnosis in the US since October 2021 with the implementation of the International Classification of Diseases (ICD) or ICD-10-CM code U09.9 for post-COVID conditions [68]. Despite this acknowledgement, treatment guidelines are still being developed [34, 35]. Long Covid can affect anyone who has had Covid-19, regardless of age, gender, or pre-existing conditions, even if the initial infection was mild [47], but may have a particular impact on older adults. It may be disproportionately diagnosed in working-age individuals, especially those aged 50-64 [48]. Factors influencing differential impact for older adults include higher Covid-19 mortality in older adults [40, 48], higher vaccination rates among older adults, data from before contemporary variants, and potential underreporting due to lower survey participation in older adults [48, 52]. Despite lower rates of documented Long Covid cases, about one in four noninstitutionalized adults over 65 have developed at least one long-term health issue following a Covid-19 infection [2]. The impact on older adults in nursing homes is not well known, but is thought to be more severe [1, 49, 50]. Compared to other long illnesses, Long Covid may cause more memory, cognitive, and sleep difficulties in older adults [52].

The varied symptoms make it difficult to determine how many of the over 650 million Covid-19 cases to date have resulted in Long Covid [1]. One in four adults aged 65 and older (in contrast to one in five younger adults) who have contracted COVID-19 have experienced at least one long-lasting health problem, based on estimates from the Centers for Disease Control and Prevention (CDC) [67]. Long Covid; referred to as post-acute sequelae of SARS-CoV-2 infection (PASC), manifests with various symptoms, such as fatigue, cognitive difficulties, headaches, breathlessness, and chest discomfort. These symptoms may continue for weeks, months, or even years following the initial infection. There is currently no specific diagnostic test, complicating the identification and management of the condition, but interim guidance and case definitions exist [51, 35, 38-40]. Long Covid is considered a disability under the Americans with Disabilities Act if it substantially limits one or more major life activities. The classification recognizes the impact of Long Covid and protects individuals from discrimination [54].

Long Covid is particularly challenging to recognize in older adults due to overlapping symptoms with other chronic diseases [1, 52]. Research shows Long Covid can cause brain abnormalities and cognitive impairments like those seen in Alzheimer’s Disease [59]. One study found that nursing home residents with Covid-19 needed increased assistance with daily activities than their uninfected peers, and this effect persisted for months [50]. Residents with Covid-19 infections and dementia experienced faster cognitive decline than peers without dementia. Residents without dementia regained their functional status nearly on par with their uninfected peers after a year. The study highlights the vital requirement for attentive management of Long COVID symptoms, especially in individuals with dementia, because of their heightened susceptibility [50]. Restoring functional status following a Covid-19 infection and careful management of Long Covid symptoms is essential in this vulnerable population particularly for at-risk groups like those with dementia who have experienced Covid-19 as they are more likely to experience severe consequences and face challenges in following preventive measures.

Without established diagnostic criteria and treatment protocols, distinguishing between Long Covid and dementia is challenging, leading to potential misdiagnoses and adverse outcomes, especially for those with existing dementia [53]. Effective care and support are crucial, alongside understanding the cause of cognitive issues for monitoring progression and meeting basic needs. Functional status, measured by Activities of Daily Living (ADLs), includes essential skills like walking, toileting, grooming, and eating. [18], Functional status is crucial in gerontological and health services research for assessing needs and independence [19, 20]. ADLs predict health outcomes [19, 20], including life expectancy, quality of life [21-23], and mortality [24], especially in patients with Alzheimer’s Disease Related Dementia [25]. Despite their importance, ADLs are often absent from clinical effectiveness literature [25, 26]. This absence is likely because ADLs are not available in claims data (a dominant data source for outcomes research for the past 25 years [66], and this data is not otherwise widely available. ADLs are used in SNFs, as mandated by the Centers for Medicare & Medicaid Services (CMS) to assess care needs and progress in older adults [59], where they are tracked within residents’ electronic health records (EHR) as often as multiple times per day.

Functional status including ADLs remain critical components in assessing the effects in the elderly population. A recent study conducted by Fésü et al. (2025) offers perspective on practical impacts of Long Covid regarding functional outcomes and patient-reported experiences. The study revealed that a significant percentage of previously hospitalized Covid-19 patients indicated ongoing symptoms and functional limitations even months following their original infection, highlighting that 60% of patients continued to show symptoms, and numerous individuals faced challenges with sleep, movement, and everyday tasks. The deterioration in functional ability among elderly individuals with Long Covid is especially alarming as it may result in greater reliance on others and a diminished quality of life. The importance of early and precise assessment of functional status is essential to inform treatment and rehabilitation strategies [61].

We describe Covid-19 treatments, functional status, hospitalization, and mortality among a population of older adults with Long Covid and other conditions including dementia.

Older individuals experiencing Long Covid frequently undergo various treatments designed to alleviate their symptoms, such as antiviral drugs like remdesivir, which have demonstrated a decrease in mortality rates independent of vaccination status. However, hospitalization rates for older adults with Long Covid continue to be elevated, especially among individuals with preexisting health issues like dementia. The elderly population make up a considerable share of hospitalizations related to Covid-19, with numerous needing intensive care and extended hospitalizations further emphasizing the need for functional status measures and providing targeted treatment.

Based on the literature above, we hypothesize that vaccinated individuals will exhibit significantly lower mortality rates compared to those who are not vaccinated. The SAVALO study demonstrated that vaccinated individuals exhibited reduced mortality rates compared to those who were not vaccinated (0.5% vs. 7.8%, p < 0.001). Nirmatrelvir/ritonavir considerably lowered mortality rates irrespective of vaccination status (0.5% in treated vs. 7.8% in untreated, p < 0.001). However, Nirmatrelvir/ritonavir solely reduced hospitalization rates for individuals who were unvaccinated. These results highlight the significance of prompt antiviral treatment in minimizing severe consequences and increasing survival rates among elderly individuals suffering from Long Covid [62].

## DATA SOURCE

We describe and analyze commercially available EHR data for 23,632 residents in nursing homes in the United States, from March 1, 2020, through June 30, 2023, diagnosed with Long Covid as identified by ICD-10-CM code U09.9. Longitudinal tracking of patient health over time allowed us to follow individuals’ health trajectories, monitor changes in health status, and acute treatments and interventions 90 days prior to and up to six months following Long Covid diagnoses. These data reflect real-world clinical practice and patient care scenarios, capturing the complexities of healthcare delivery in an important clinical setting.

This study was conducted in accordance with the ethical standards set forth by the Jefferson College of Population Health Thomas Jefferson University Institutional Review Board (IRB) and the Declaration of Helsinki. The IRB issued a waiver of exemption for this study. The anonymity of all data was maintained throughout the study.

## MEASURES

| Data Element | Description |
| --- | --- |
| Functional Status (baseline) | The mean of each resident's ADL scores (scores range from 0-16 and indicate how well individuals can perform daily tasks such as bathing, dressing, toileting, transferring, continence, and eating [12]) in the two weeks prior to onset of Long Covid. Higher ADL scores indicate more dependence on caregivers; lower ADL scores indicate more independence in physical function. |
| Function Status (outcome) | The mean of each resident's ADL scores over a two-week window, six months following the onset of Long Covid. For logit models, we use a binary measure of ADL change as either stable/better (ADL scores the same or lower than at Long Covid onset) or worse (ADL scores higher in the six months after Long Covid onset). |
| Demographic Attributes | Average age and % male |
| Mortality Rate | The percentage of deaths in the cohort within six months following Long Covid onset. |
| Hospitalization Rate | The percentage of residents with at least one hospital visit within 6 months following Long Covid onset. |
| Covid Risk Factors | The average number of risk factors (total number of diagnosed conditions prior to Long Covid onset; identified by ICD-10 diagnosis codes) was included; Dementia and Chronic Kidney Disease were included additionally as individual variables.<br>A study released in JAMA Internal Medicine found that cognitive impairment, such as dementia, was strongly linked to higher mortality rates among Covid-19 patients, emphasizing the susceptibility of this group [64].<br>A study in BMC Geriatrics identified a significant link between Covid-19 infection and the emergence of new-onset dementia in elderly individuals, highlighting the prolonged neurological dangers posed by the virus [63]. |
| Body Mass Index (BMI) | A flag for BMI > 25 was included; In this population, BMI greater than 25 may be protective against certain conditions and may be associated with better health outcomes (standard BMI ranges do not necessarily apply to older adults due to changes in body composition, such as decreased muscle mass). |
| Vaccination Status | A flag for whether a resident had any documented Covid-19 vaccine prior to the onset of Long Covid |
| CMS Five-Star Ratings | The CMS Five-Star Quality Rating System is a tool created by the CMS features a quality rating system that assigns each nursing home a rating from 1 to 5 stars, with 5 stars indicating much above average quality. The ratings are based on three domains: health inspections, staffing, and quality measures [58]. |
| Covid-19 Treatments | Treatments for acute Covid-19 (Covid-19) have included a great variety of medications like Paxlovid, Remdesivir, and monoclonal therapies like Bebtelovimab, Sotrovimab, Tixagevimab & Cilgavimab. |

Vaccination for Covid-19 prior to Long Covid onset is included in all models. Table 1 presents more detail on vaccination across population and treatment groups.

**Table 1.** Demographic, Clinical, and Facility Characteristics Among Long Covid Cases by Treatment Received.

| Treatment | No treatment | Sotrovimab | Paxlovid | Molnupiravir | Bebtelovimab | Remdesivir | Tixagevimab & Cilgavimab |
| --- | --- | --- | --- | --- | --- | --- | --- |
| N = 23,653 | 21,815 | 40 | 1123 | 340 | 21 | 9 | 1 |
| % | 93.43% | 0.17% | 4.81% | 1.46% | 0.09% | 0.04% | 0.00% |
| Average Age | 76.8 | 78.6 | 80.0 | 78.5 | 77.9 | 82.0 | 77.0 |
| Percent Male | 47% | 33% | 39% | 39% | 38% | 56% | 100% |
| Average Risk Factors | 3.3 | 3.8 | 3.2 | 3.6 | 3.3 | 3.2 | 0.0 |
| % with Dementia | 27% | 50% | 41% | 37% | 55% | 56% |  |
| % with CKD | 32% | 40% | 29% | 44% | 25% | 22% |  |
| % White | 65% | 78% | 72% | 79% | 67% | 67% | 100% |
| % with vaccine before onset | 56% | 93% | 80% | 83% | 81% | 78% | 100% |
| Avg ADL | 5.1 | 3.8 | 5.9 | 6.4 | 9.7 | 12.7 | 5.0 |
| Facility Five-Star Overall Rating | 3.0 | 3.4 | 3.1 | 3.2 | 3.6 | 3.6 | 5.0 |
| Facility Five-Star Quality Rating | 3.9 | 4.3 | 3.9 | 3.8 | 3.8 | 4.6 | 5.0 |
| Facility Five-Star Staffing Rating | 2.7 | 2.7 | 2.8 | 2.6 | 2.9 | 3.3 | 5.0 |
| Facility Five-Star Survey Rating | 2.7 | 2.8 | 2.9 | 3.0 | 3.6 | 2.6 | 5.0 |

## ANALYSES

We describe pharmacotherapy across US residents within SNFs across patient-level factors including demographics, comorbidities/risk factors, and facility quality. Most individuals diagnosed with Long Covid did not receive included treatments. The group with no treatments had the lowest mean age (76.8 years), was composed of 46% males, and had an average baseline ADL score of 5.1, indicating moderate functional limitations. The group that received Remdesivir is the oldest and most functionally limited with a mean age of 82 years and average baseline ADL score of 12.7. The group that received Paxlovid has the lowest average number of risk factors (equal to that of the Remdesivir group), a relatively low percentage of people diagnosed with dementia at 41%, and 80% of this group were vaccinated for Covid-19 compared to 56% of the group with no treatment.

“Block” regression models illustrate covariance and spurious relationships between variables. Rows show how coefficients change when other variables are included in models (e.g. Coefficients in Model 1 may be altered by including additional variables in Model 2, with which they covary.) The “Block” models are therefore not evaluated for fit but used for descriptive purposes. For each outcome, Model 5 contains drug treatment groups and all control variables.

### Mortality (Table 2a)

Remdesivir has the biggest association with lower mortality risk compared to No Treatment, controlling for variables such as age, functional status, and number of co-occurring conditions. These variables each have statistically significant coefficients smaller than -0.02. Table 1 shows that the Remdesivir group is older and has worse functional status on average, which mathematically makes it harder for a drug to show improvement; Model 5 shows how age, treatment, and mortality are related to factors such as dementia which significantly increases mortality risk, number of co-occurring conditions which slightly reduces risk (potentially capturing access to specialized care), functional status as expected given less room for improvement among those with high ADL scores, and facility quality and vaccine status significantly reducing risk. Paxlovid is associated with a non-significant reduced risk of mortality.

**Table 2:** Block Regression Model Outcomes for Long Covid Treatment Groups.

| Model | Age | Condition count | CKD | Dementia | BMI > 25 | Female Sex | ADL | Vaccine | Five-Star Rating | Bebtelovimab | Molnupiravir | Paxlovid | Remdesivir | Sotrovimab |
| --- | --- | --- | --- | --- | --- | --- | --- | --- | --- | --- | --- | --- | --- | --- |
| Model 1 | -0.03** | -0.11** | 0.41** | 0.59** | -0.72** | -0.34** |  |  |  |  |  |  |  |  |
| Model 2 | -0.03** | -0.11** | 0.39** | 0.64** | -0.72** | -0.32** | -0.1** |  |  |  |  |  |  |  |
| Model 3 | -0.03** | -0.11** | 0.4** | 0.65** | -0.72** | -0.32** | -0.09** | -0.16** |  |  |  |  |  |  |
| Model 4 | -0.03** | -0.1** | 0.38** | 0.61** | -0.71** | -0.31** | -0.09** | -0.14** | -0.12** |  |  |  |  |  |
| Model 5 | -0.02** | -0.1** | 0.38** | 0.62** | -0.71** | -0.31** | -0.09** | -0.13* | -0.12** | -15.44 | 0.07 | -0.21 | 2.28** | -0.19 |

**B) Hospitalization within six months of Long Covid onset**
| Model | Age | Condition count | CKD | Dementia | BMI > 25 | Female Sex | ADL | Vaccine | Five-Star Rating | Bebtelovimab | Molnupiravir | Paxlovid | Remdesivir | Sotrovimab |
| --- | --- | --- | --- | --- | --- | --- | --- | --- | --- | --- | --- | --- | --- | --- |
| Model 1 | -0.02** | 0.1** | 0.3** | -0.2** | -0.25** | -0.16** |  |  |  |  |  |  |  |  |
| Model 2 | -0.02** | 0.1** | 0.3** | -0.2** | -0.25** | -0.16** | 0 |  |  |  |  |  |  |  |
| Model 3 | -0.02** | 0.1** | 0.3** | -0.2** | -0.25** | -0.16** | 0 | -0.04 |  |  |  |  |  |  |
| Model 4 | -0.02** | 0.11** | 0.3** | -0.21** | -0.25** | -0.15** | 0 | -0.03 | -0.06** |  |  |  |  |  |
| Model 5 | -0.02** | 0.11** | 0.3** | -0.21** | -0.24** | -0.15** | 0 | -0.02 | -0.06** | 0.45 | -0.23 | -0.21* | 0.22 | 0.03 |

**C) Functional status decomposition six months after Long Covid onset**
| Model | Age | Condition count | CKD | Dementia | BMI > 25 | Female Sex | ADL | Vaccine | Five-Star Rating | Bebtelovimab | Molnupiravir | Paxlovid | Remdesivir | Sotrovimab |
| --- | --- | --- | --- | --- | --- | --- | --- | --- | --- | --- | --- | --- | --- | --- |
| Model 1 | -0.06** | -0.09** | 0.26* | 1.77** | -0.41* | 0.1 |  |  |  |  |  |  |  |  |
| Model 2 | -0.06** | -0.11** | 0.35** | 1.54** | -0.41* | 0.03 | 0.17** |  |  |  |  |  |  |  |
| Model 3 | -0.07** | -0.14** | 0.37** | 1.52** | -0.4* | 0.03 | 0.15** | 1.04** |  |  |  |  |  |  |
| Model 4 | -0.07** | -0.13** | 0.36** | 1.49** | -0.39* | 0.04 | 0.15** | 1.07** | -0.09* |  |  |  |  |  |
| Model 5 | -0.07** | -0.12** | 0.32* | 1.61** | -0.47** | 0.03 | 0.14** | 1.15** | -0.13** | -0.03 | -0.25 | -0.52* | -0.01 | 0.21 |

We describe relationships between patient-level factors and pharmacotherapy indicators against three separate patient outcomes: mortality, rehospitalization, and decomposition in functional status.

### Hospitalization (Table 2b)

Paxlovid is associated with a significantly reduced risk of hospitalization, along with protective factors like having a BMI greater than 25, being female, and hypothetically having specialized care due to conditions like dementia and kidney failure. Facility quality also slightly reduced the risk of hospitalization. Perhaps due to high variability in complications for older adults with Long Covid and other terms in Models 2-5, ADLs capture virtually zero variation relevant to hospitalization.

### Decompensation of Functional Status (Table 2c)

Paxlovid is strongly and significantly associated with reduced risk of functional decompensation. To a lesser extent, having a BMI greater than 25 and, hypothetically, having specialized care due to conditions like dementia and kidney failure may also be protective factors. Facility quality also slightly reduced risk of decompensation as expected; risk of decompensation based on ADL score at baseline appears mitigated, at least in part, by care provided as captured by vaccination (accounts for a -0.02 change in ADL coefficient) and treatment with medication (accounts for a -0.03 change in ADL coefficient).

Paxlovid appears with the strongest protective association against decompensation. This is noteworthy, as in review of 18 randomized clinical trials or observational trials of adult Covid-19 survivors, all 18 studies found a worsening ADL performance and consequent loss of independence in patients after the acute phase of infection [55]. Case reports [11, 12] involving Long Covid patients whose symptoms improved following Nirmatrelvir-Ritonavir also align with our findings. One study with 82% vaccinated nonhospitalized US veterans used EHR data to examine how Paxlovid affects post-Covid outcomes. It was found that Paxlovid and Molnupiravir both associated with reduced hospitalization and mortality risk at 30 days and six months [9], both treatments working better than no treatment, and no difference was seen between them.

### Vaccination

Tables 2a. highlights vaccination as a significant potential protective factor against mortality for all groups, including and possibly especially for the No Treatment group (Table 2a Models 3-5). Vaccination for Covid-19 may be associated with a lower likelihood of mortality from Long Covid due to factors including the reduction of viral load in vaccinated individuals, protection against variants of concern, modulation of the immune response to regulate inflammation associated with Long Covid, and secondary effects on overall health such as mental well-being and healthcare-seeking behavior. Vaccination campaigns have been linked to improvements in mental health, increased healthcare-seeking behavior, and better adherence to preventive measures, all of which may indirectly contribute to a lower likelihood of mortality from Long Covid. However, relationships for hospitalization are non-significant (Table 2b).

For functional limitation outcomes, the relationships are reversed: in all models and particularly for the No Treatment group, vaccination is a significant predictive factor for decompensation (Table 2c).

## DISCUSSION

Fyffe et al. identified four key factors that improved outcomes for Long Covid survivors in long-term care: awareness, accountability, vigilance, and empathetic listening [33]. As Long Covid can cause anxiety, depression, and social isolation, clinicians can help residents cope with their mental health and recovery.

Data and observations from long-term care can help understand Long Covid and its effects on function, which can guide further research and interventions.

The study explores the relationships between patient-level factors, pharmacotherapy indicators, and three patient outcomes: mortality, rehospitalization, and functional status decompensation. The use of “block” regression models helps illustrate how coefficients change when additional variables are included, providing a descriptive rather than a fit-evaluated analysis. These descriptions suggest that pharmacotherapy, particularly with Remdesivir and Paxlovid, along with vaccination and high-quality care, can significantly improve outcomes for older adults in nursing homes with Long Covid.

### Strengths & Limitations

The current study does not account for time dependence. Exploring time-dependent factors and expanded indicators of selection into treatment groups and vaccine status may help suggest mechanisms for our findings above. For example, more vulnerable older adults may have selected into the vaccination group earlier, and this group may have increased propensity for decompensation. Decompensation associated with vaccination may also help identify types of individuals for whom vaccination is nonprotective versus those for whom vaccination is likely protective. We also pool all people with vaccines prior to Long Covid onset and do not compare vaccine types (e.g. mRNA vaccines) or brands [29], and vaccination after onset was not explored. Another limitation is our use of ICD-10-CM code U09.9 as the sole criterion for defining Long Covid, which potentially biases the population as inclusion is dependent on receiving a formal (and documented) diagnosis. Since numerous individuals with Long Covid are never officially diagnosed, the population of individuals including in our study may not be representative of the Long Covid population as a whole. Future research might gain from focusing more on symptoms/observations instead of depending exclusively on ICD-10 diagnoses. This method prepares for the subsequent study to employ observational data, possibly offering a deeper insight into Long Covid.

Analyzing additional conditions, medications, assessments, observations, and symptoms could help create a profile of Long Covid in older adults in care facilities and ways to treat and manage it. For example, David et al surveyed people with Covid-19 and found an average of 55.9+/-25.5 symptoms in 9.1 organs. They identified 203 symptoms in 10 organs and tracked 66 symptoms for seven months for 3,762 people. Doing a similar analysis for older adults, especially those with cognitive problems like dementia and social challenges like documented barriers to health care, could help compare adult and older adult groups and better define Long Covid. An article by Mandal et al. revealed that systematic follow-up after hospitalization for COVID-19 tracked the progression of physical and psychological symptom burden, recovery of blood biomarkers, and imaging irregularities. This method may be especially advantageous for elderly individuals in care homes, offering insights into the need for rehabilitation and further study, allowing this detailed profiling and subsequent follow-up to guide tailored interventions and enhance management approaches for Long COVID in this risk group [65].

During the preparation of this work the author(s) used Microsoft Word 365 to improve readability. After using this tool/service, the author(s) reviewed and edited the content as needed and take(s) full responsibility for the content of the publication.

## Acknowledgements

No funding was received for this study.

## Notes

### Competing Interest Statement

The authors have declared no competing interest.

